# Optimizing UK Biobank Cloud Based Research Analysis Platform to Fine Map Coronary Artery Disease Loci in Whole Genome Sequencing Data

**DOI:** 10.1101/2024.09.23.24313932

**Authors:** Letitia M.F. Sng, Anubhav Kaphle, Mitchell J. O’Brien, Brendan Hosking, Roc Reguant, Johan Verjans, Yatish Jain, Natalie A. Twine, Denis C. Bauer

**Affiliations:** Australian e-Health Research Centre, Commonwealth Scientific and Industrial Research Organisation (CSIRO), Westmead, New South Wales, Australia; Australian e-Health Research Centre, Commonwealth Scientific and Industrial Research Organisation (CSIRO), Melbourne, Victoria, Australia; Australian institute for Machine Learning, University of Adelaide, Adelaide, South Australia, Australia; Lifelong Health, South Australian Health and Medical Research Institute, Adelaide, South Australia, Australia; Royal Adelaide Hospital, Central Adelaide Health Network, Adelaide, South Australia, Australia; Applied BioSciences, Faculty of Science and Engineering, Macquarie University, Macquarie Park, New South Wales, Australia; Australian e-Health Research Centre, Commonwealth Scientific and Industrial Research Organisation (CSIRO), Adelaide, South Australia, Australia; University of Sydney, School - School of Medical Sciences, Department of Biomedical Informatics and Digital Health, Sydney, Australia

**Keywords:** Population-Scale Genetics, UKBiobank, DNAnexus, Cloud-computing, GWAS, Trusted Research Environments

## Abstract

We conducted the first comprehensive association analysis of a coronary artery disease (CAD) cohort within the recently released UK Biobank (UKB) whole genome sequencing dataset. We employed fine mapping tool PolyFun and pinpoint *rs*10757274 as the most likely causal SNV within the 9p21.3 CAD risk locus. Notably, we show that machine-learning (ML) approaches, REGENIE and VariantSpark, exhibited greater sensitivity compared to traditional single-SNV logistic regression, uncovering *rs*28451064 a known risk locus in 21q22.11. Our findings underscore the utility of leveraging advanced computational techniques and cloud-based resources for mega-biobank analyses. Aligning with the paradigm shift of bringing compute to data, we demonstrate a 44% cost reduction and 94% speedup through compute architecture optimisation on UK Biobank’s Research Analysis Platform using our RAPpoet approach. We discuss three considerations for researchers implementing novel workflows for datasets hosted on cloud-platforms, to pave the way for harnessing mega-biobank-sized data through scalable, cost-effective cloud computing solutions.

## Introduction

In November 2023, the UK Biobank (UKB) released the world’s largest single set of whole genome sequencing (WGS) data of half a million individuals through their Research Analysis Platform (RAP). In total, 27.5 petabytes of data was created as outlined by UK Biobank WGS consortium [1]. Briefly, the GraphTyper-called [2] WGS data comprised of 1,037,556,156 SNVs and 101,188,713 indels across the 490,640 participants, resulting in 1510 terabytes across 145,207 pVCFs (Supplementary Table S1).

Implementing a new paradigm of data management, the data is stored on the RAP, a cloud-based platform, which allows analysis workflows to be “brought to the data”. Registered RAP users with a linked UKB Access Management System (AMS) account can access and analyse datasets through “applets”, including the Swiss Army Knife (SAK) app, which holds several bioinformatics tools such as PLINK2 [3] and bcftools [4] as well as user-installable tools for custom workflows. Besides the Tier 3 data access fee to UKB, other costs to the user via DNAnexus include storing uploaded or derived data, egress, and compute using Amazon Web Services (AWS).

The use of such cloud-based Trusted Research Environments (TREs) is becoming more commonplace as genomic studies increase in scale (e.g., *All of Us* [5]). While these TREs offer benefits like enabling cross-cohort analysis [6], they also present distinct challenges, including a steep learning curve for researchers unfamiliar with cloud and bioinformatics, and costs that scale with cohort size. Given the size of the UKB WGS dataset, the costs to run a genome-wide association analysis would be substantial, particularly if compute is not optimised. Therefore, this study set out to explore the usability of the RAP and the potential cost of running an association analysis on the UKB WGS dataset.

We present the first coronary artery disease (CAD) association analysis that leverages the unprecedented density of the UKB WGS dataset facilitated by our parallelisation tool, RAPpoet, and show how the density of the dataset enables novel insights into CAD genetics. We also discuss how bringing the compute to the data facilitates individual-level access for researchers worldwide and three considerations for researchers looking to use cloud-based TREs.

## Results

### RAPpoet: A Driver-Worker Approach to Running Jobs on the UKB RAP

As of writing, the web UI of RAP and available tutorials are tailored towards processing UKB’s genomic data sequentially or in small batches. While this was well suited to earlier and smaller versions of the UKB datasets (e.g., array imputed data), with 145,207 pVCFs this approach has become impractical.

To address this, we have designed a scalable orchestration engine called RAP parallelisation orchestration engine template (RAPpoet), to streamline our pipeline across the UKB pVCFs using the DNAnexus CLI (Figure 1).

**Figure 1.**
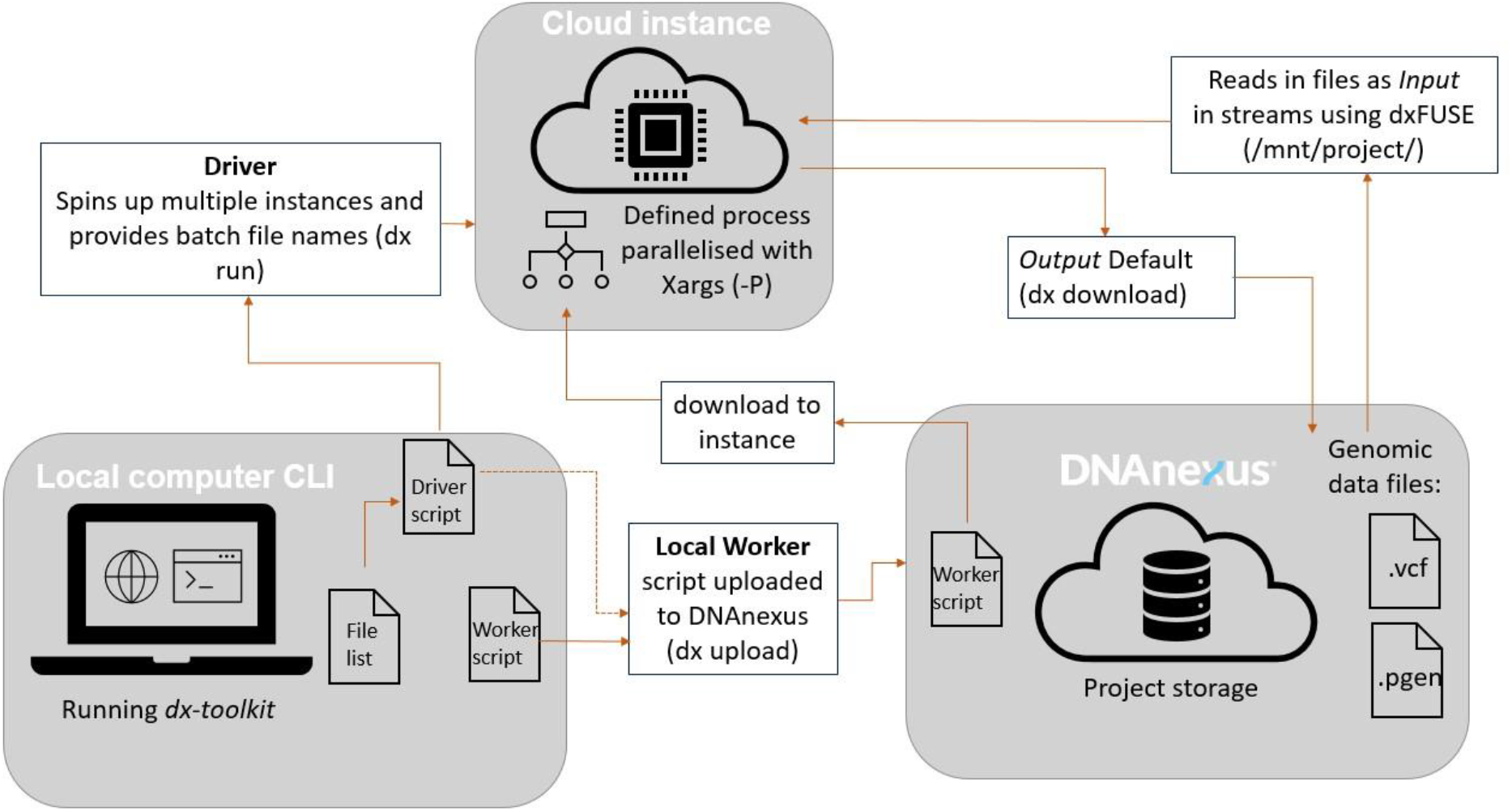
A Driver-Worker Approach (RAPpoet) for Managing the Configuration of Requested Instances on UK Biobank RAP and for Parallelising the Processing of Files on Cloud Instances. The driver script, which remains on the user’s local machine, includes configurable environmental variables, such as the type of instance requested and number of batches to process in each instance. Meanwhile, the worker script, located in the RAP project environment, contains the actual commands to be executed on the instance that can be set by the user depending on their requirements.

RAPpoet utilises two scripts: the ‘driver’ and the ‘worker’. The ‘driver’ script sits within the user’s local compute and includes commands for configuring the instance environment, uploading required files, and initiating the execution of the ‘worker’ scripts on requested instances. The ‘worker’ script is uploaded onto each instance and defines the processes to be run over the uploaded files.

This setup enables the parallelisation of tasks as the ‘worker’ script can execute the defined process concurrently via the xargs tool, optimising resource utilisation by getting a single instance to handle multiple files. This also decreases the number of instances needed making it easier to manage and monitor jobs.

Additionally, RAPpoet streamlines the workflow by centralising the control and coordination in the ‘driver’ script. This improves the overall manageability of jobs by consolidating file setup and instance configuration within the ‘driver’ script. Essential variables, such as instance types, file lists, batch sizes, and the number of files run in parallel, are defined, ensuring consistency across all instances. Furthermore, centralising control to the ‘driver’ script mitigates potential issues related to file duplication and ensures a more organised and streamlined execution.

Finally, RAPpoet facilitates efficient multi-tasking. While the RAP’s web UI typically executes one command at a time on uploaded files, our approach allows the user to define a more comprehensive process within the ‘worker’ script, such as running multiple bcftool commands in a single job. This capability also enables the execution of multiple steps calling on different tools within a single job, further enhancing the flexibility and efficiency of each task.

### Optimising Cloud Compute and Parallelisation is Crucial to Reducing Cost and Runtime on RAP

On the RAP, analyses are carried out on AWS Elastic Compute Cloud (EC2) instances with different options for storage, memory capacity, and number of cores. In this study, we evaluated the impact of different compute architectures and of running files in parallel versus sequentially on cost and runtime, particularly for the requisite quality control (QC) steps.

Running QC step one (Figure 5) on a single chromosome 21 pVCF of 10.5GB, the average pVCF size, using the RAP’s Web UI and Workflow approach took 30 minutes. In contrast, using RAPpoet to parallelise across multiple files, the average runtime for one pVCF was 1.75 minutes (Supplementary Table S10), a 94% reduction in runtime compared to the sequential approach. Running all 1867 pVCF in Chromosome 21 and 6106 pVCF in Chromosome 9 using RAPpoet’s parallelisation approach, runtime for this step was 323 minutes and 1103 minutes respectively (Table 1).

**Table 1.** Summary of Cost and Runtime for Association Analysis Pipeline using RAPpoet. Note that the cost and runtime of each step is a function of EC2 instance type, job priority, etc. See Supplementary Material and Tables for more detailed costings and runtime. Note that fine mapping was run on a locally as PolyFun was not available on the UKB RAP.

| Step | Chromosome 9 |  | Chromosome 21 |  |
| --- | --- | --- | --- | --- |
|  | Cost (£) | Runtime (Min) | Cost (£) | Runtime (Min) |
| Quality Control Step One | 176.50 | 1103 | 97.05 | 323 |
| Quality Control Step Two | 33.33 | 855 | 5.41 | 216 |
| sLR | 0.078 | 15 | 0.033 | 6 |
| REGENIE | 2.75 | 91 | 0.23 | 31 |
| VariantSpark | 179.31 | 796 | 56.14 | 499 |

Furthermore, by optimising compute instances, the cost associated with QC step one of the pipeline (***Figure 5***) was reduced by 44% (£0.029 per file vs £0.052) between chromosome 9 and 21 while keeping runtime constant. Initially, the instance type mem1_ssd1_v2_x72 was selected resulting in an average runtime of 1.75 minutes and an average cost of £0.052 per pVCF (Supplementary Table S10). However, by switching to the smaller instance mem2_ssd1_v2_x48 type (i.e., half the number of vCPUs), the average runtime and cost per pVCF for QC step one on chromosome 9 was 1.80 minutes and £0.029 (Supplementary Table S11).

Other settings that impact cost and runtimes such as job priority and file I/O were also benchmarked (Supplementary Material S1). Notably, the dxFUSE I/O filesystem limits the number of files that can be processed concurrently, severely hindering the level of parallelisation of RAPpoet and requiring a looping system within RAPpoet to process the 6,106 pVCFs from chromosome 9.

### Machine Learning Methods Show Greater Potential Than Logistic Regression for High Density Genomic Data

Although this finding is secondary to the focus of this study, the association analysis pipeline, from QC to fine mapping using RAPpoet, revealed interesting trends. In particular, how ML approaches compared to the traditionally used single-SNV logistic regression (sLR).

The significance of the well-established 9p21.3 (*CDKN2B-AS1*) risk locus [7,8] was confirmed with sLR (Figure 2, Supplementary Table S2). The unprecedented density of the UKB WGS dataset further enabled fine mapping to identify the most likely causal variant. The same 13 SNVs were identified in the first credible sets of both FINEMAP [9] and SuSie [10] (Figure 3; Supplementary Tables S3 and S4). Of these, *rs*10757274 has been linked to *CDKN2B-AS1* expression [11,12], is part of the core risk haplotype region [13], and has been associated with CAD in several populations [14,15], suggesting that *rs*10757274 is the most likely causative SNV in the risk locus.

**Figure 2.**
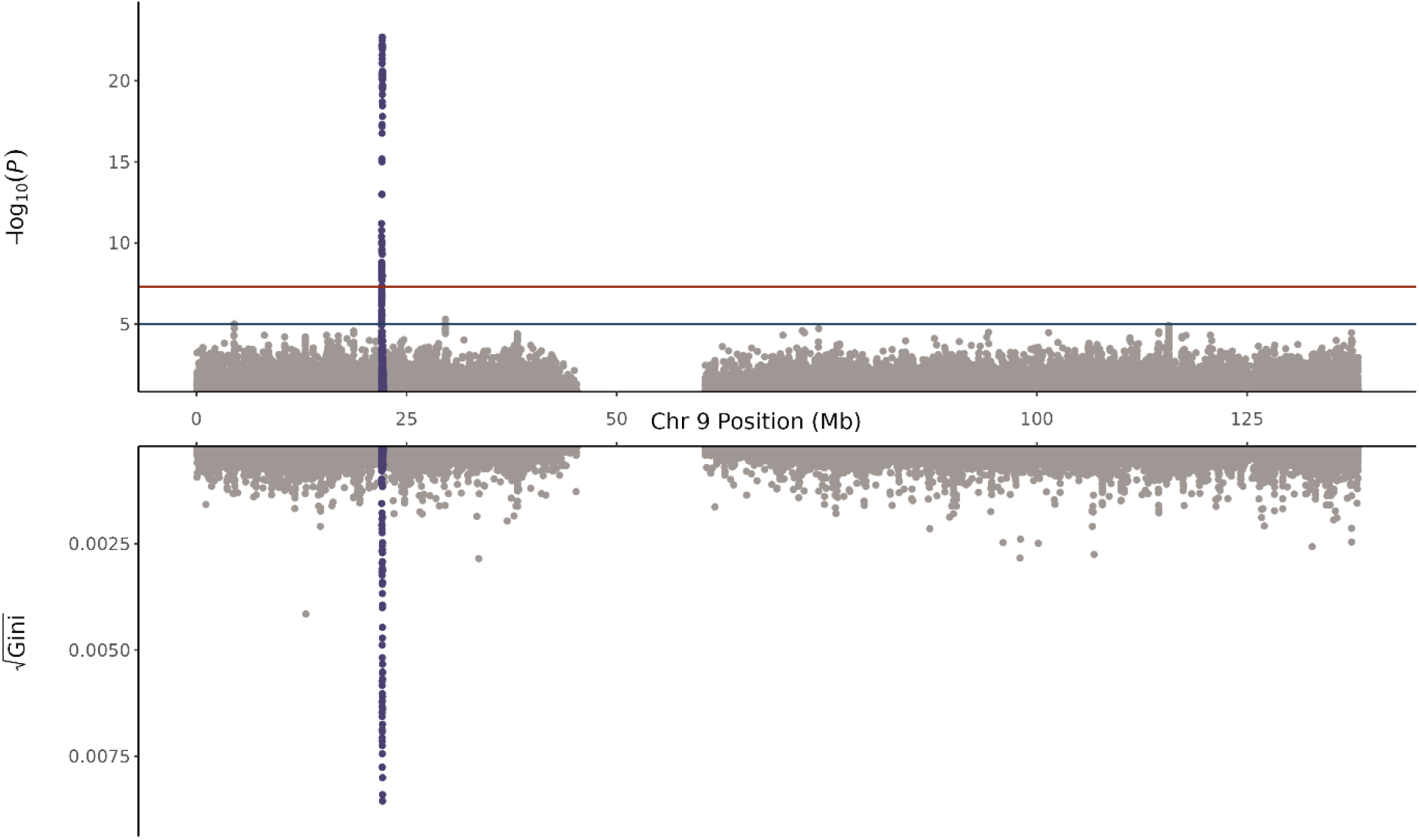
Miami Plot of Chromosome 9 for single-SNV Logistic Regression (Top) and VariantSpark (Bottom). The locus 9p21.3 reached genome-wide significance in the logistic regression analysis and is the top association from VariantSpark. Note that the Gini score (bottom *y*-axis) is used to measure variable importance and while it is analogous to the traditional P-values of logistic regression to rank associations relative to other associations, it does not include multiple testing and does not have guiding significance thresholds.

**Figure 3.**
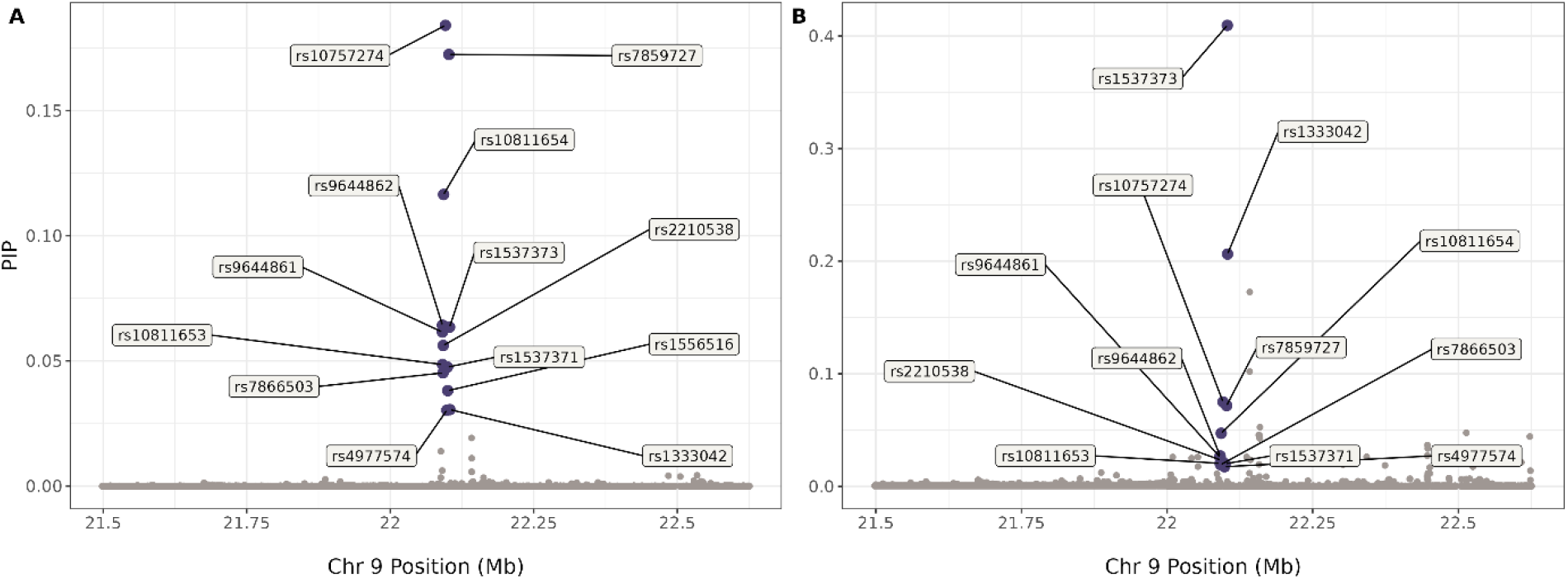
Scatterplot of SNVs in the 9p21.3 locus with Posterior Inclusion Probability (PIP) (y-axis) against Chromosome 9 Position (x-axis) for FINEMAP (A) and SuSie (B). The first credible set of both approaches include the same 13 SNVs which are highlighted in dark purple and annotated with *rs*IDs.

The same region and *rs*10757274 were also identified using the ML GWAS platforms VariantSpark [16] (Figure 2; Supplementary Table S5) and REGENIE [17] (Supplementary Table S6).

To further test the differences between sLR and ML on this richer data resource, the same association pipeline was also applied to chromosome 21. Figure 4 shows that sLR did not find any significant genome-wide SNVs (Supplementary Table S7). However, REGENIE found one significant genome-wide SNV, *rs*28451064 (Supplementary Table S8) in the known CAD locus 21q22.11 (*KCNE2*) [18] and VariantSpark also identifies this SNV as the top association (Supplementary Table S9), demonstrating that these ML-based approaches were more sensitive than sLR in this analysis. The *rs*28451064 SNV is intergenic between long non-coding RNA (lncRNA), *LINC00310* and the *KCNE2* gene and has been previously associated with CAD as a putative functional SNV [19,20].

**Figure 4.**
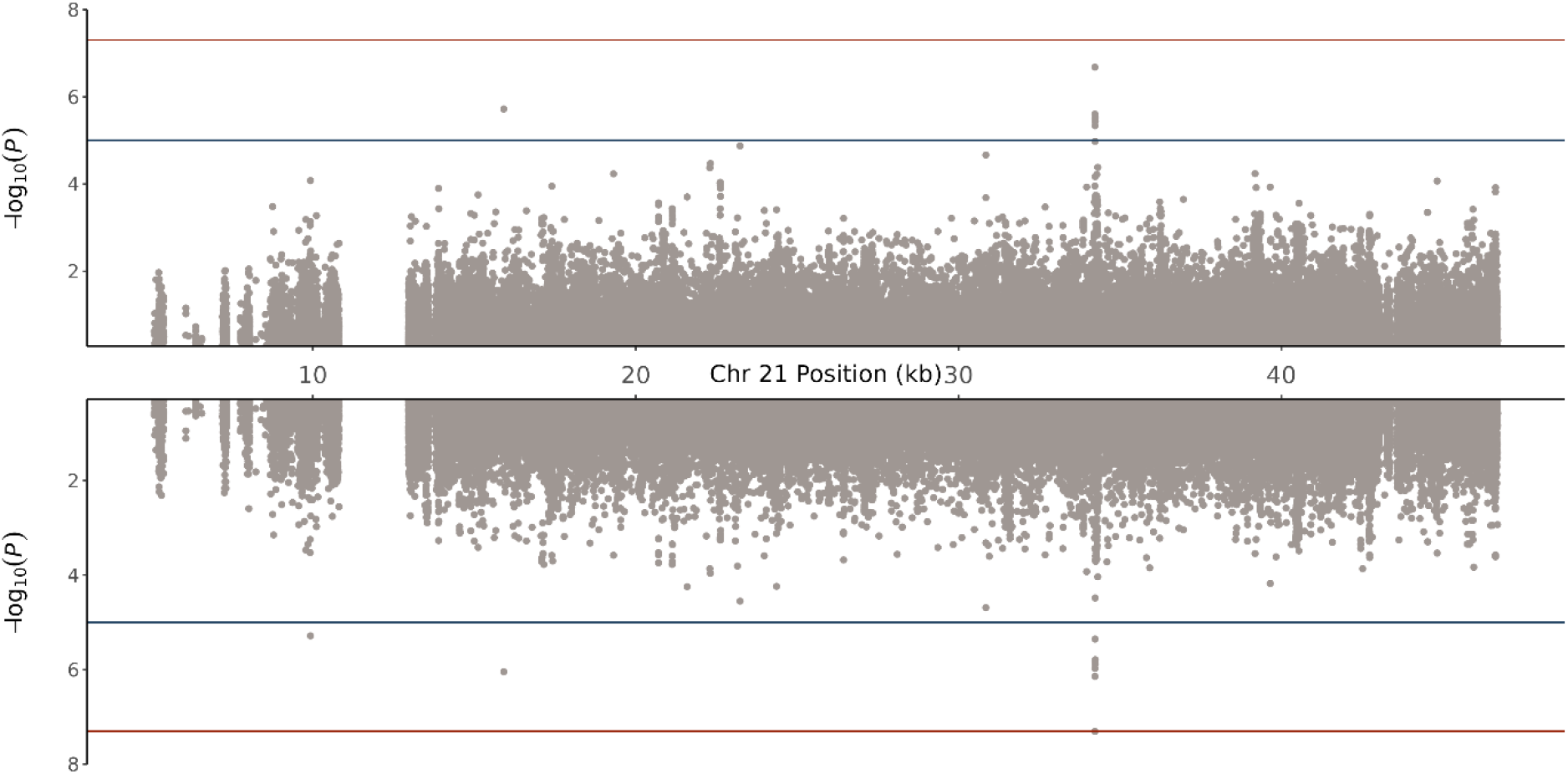
Miami Plot of Chromosome 21 for Logistic Regression (Top) and REGENIE (Bottom). The locus 21q22.11, which has been previously associated with CAD, is only showing marginal association in the single-SNV logistic regression analysis. Conversely, a single SNV in the locus reached genome-wide significance in the REGENIE analysis.

Taken together, these results highlight that the era of ML in genomics could be ushered in by the unprecedented data volumes, both in cohort size and density, that UKB-style mega-biobanks offer, potentially leading to novel findings that traditional statistical approaches may overlook. However, this finding will need to be replicated in future research across multiple diseases and/or phenotypes.

## Discussion

UKB’s RAP and DNAnexus have pioneered access to large-scale individual-level data by overcoming the risks of moving data, such as corruption, inability to enact consent changes, and lack of standardisation. In this study, we demonstrated that access to such dense individual-level data enables ML approaches to identify associations that traditional statistical methods may overlook. Additionally, previous research has highlighted potential differences in findings between individual-level analysis and meta-analysis, particularly for diverse populations [6].

Supporting this paradigm-shift, we discuss three considerations for researchers to efficiently process the UKB’s resource on RAP and other such datasets (e.g., All Of Us Researcher Workbench).

**Firstly, parallelisation is crucial** for efficient computation over the large number of files in mega-biobank-scale WGS cohorts. To streamline this process on the RAP, we designed RAPpoet. Akin to high-performance compute scheduling systems, this worker-driver architecture allows massively parallel workloads while minimising monitoring overhead (centralised coordination). It also enables multitasking within instances, thereby optimising the utilisation of compute resources. In this study, we show that using RAPpoet decreased runtime by 94% (1.75 minutes vs 30 minutes).

The upcoming inclusion of pre-processed PLINK and BGEN format files in the RAP may collapse individual pVCF files into larger cohort files. While there may be fewer files, the data volume may exhaust compute resources and still requires these monolithic files to be split into distributed workloads, which can be done with RAPpoet. Furthermore, as RAPpoet was built specifically for the RAP, if other cloud platforms such as Google Cloud become available on the RAP, RAPpoet is extendable to them. Similarly, if other biobanks that are enabled by DNAnexus follow a similar set up to RAP, RAPpoet could potentially be applicable to these data sources as well. While the architectural design of RAPpoet (i.e., the driver-worker set up) can be readily adapted to other platforms, its functionality depends on key RAP components such as the dx-toolkit which needs to be supported for RAPpoet to perform effectively.

**Secondly, cloud compute architecture needs to be tuned** to the workload, file size and time constraints at hand. On the RAP, analyses are carried out on AWS Elastic Compute Cloud (EC2) instances with different options for storage, memory capacity, and number of cores.

In this study, we show that by optimising compute resources, costs can be reduced by 44% (£0.052 per file vs £0.029) while keeping runtime constant. Furthermore, RAPpoet intrinsically optimises compute by processing multiple files concurrently rather than sequentially as suggested by the RAP tutorials. If a single pVCF was run through QC steps one and two of the pipeline (***Figure 5***) using the RAP’s Web UI and workflow approach, runtime would increase by 28.25 minutes (230% per pVCF more than RAPpoet) at minimum. Although, with the requisite skills and experience, parallelisation can be achieved through the RAP’s CLI as well [21].

**Figure 5.**
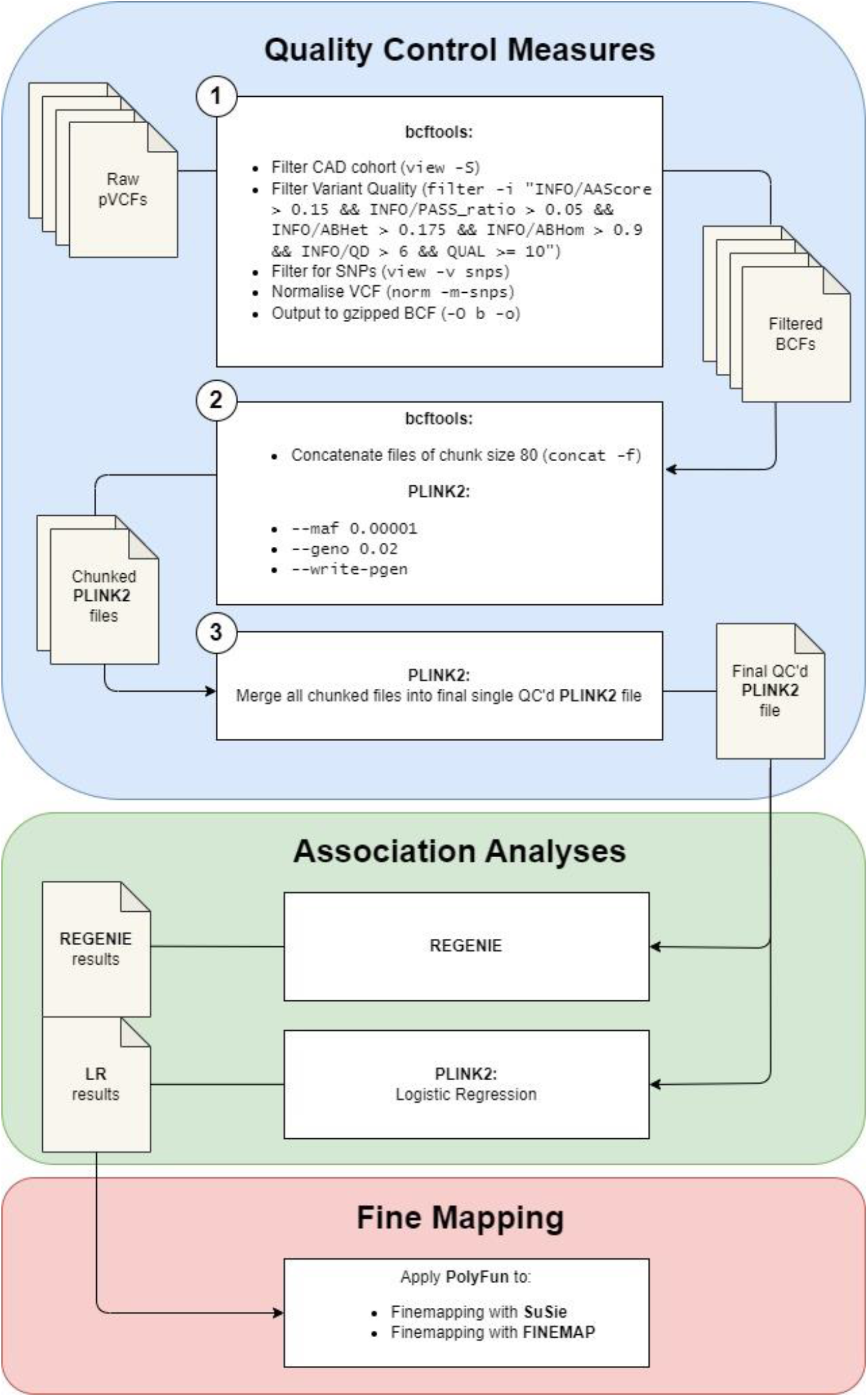
Schematic of Pipeline for UK Biobank 500K Whole Genome Sequencing Dataset on RAP. RAPpoet was used to facilitate this pipeline on the UKB RAP.

We only ran chromosome 9 and 21 in this study, as whole genome analysis is estimated to take 4.3K hours and cost £4.2K. This is due to the DNAnexus dxFUSE I/O system, which limits the number of files that can be read into the compute instances in parallel. Updates to dxFUSE are necessary to reduce the estimated runtime.

Another important factor of cost optimisation is the risk of information loss when using ‘spot’ instances (low-priority tasks), which are cheaper but have the risk of external termination compared to the more expensive ‘on-demand’ instances (high-priority tasks) with full availability (Supplementary Material S1). Workflows wanting to use low-priority tasks need to implement strategies such as checkpointing to capture machine images, which allows for the process to resume from the last captured state before termination.

**Lastly, data privacy and security must be balanced with the benefits** of making individual-level data accessible to researchers, as demonstrated in this study. Implementing federated access, including appropriate governance and seamless integration of dynamic consent layers, requires design-choices from conception [22]. Ideally such systems also enable interactive queries across genomes and metadata without full access to the raw data, such as queries enabled through sBeacon, the cloud-based implementation of the GA4GH Beacon protocol [23].

In conclusion, this study demonstrates that ML approaches are more sensitive in detecting associations from dense mega-biobank datasets compared to LR. As the field continues to adopt cloud-based platforms like the RAP to exclusively manage access to large-scale datasets, their responsibility will be to equitably support researchers in conducting innovative and technologically complex genomic studies.

## Methods

### Cohort Determination

Participants were determined to have coronary artery disease (CAD) using International Classification of Disease (ICD)-9, ICD-10, and OPCS Classification of Interventions and Procedures (OPCS)-4 codes (i.e., 410, 411, 413, 414, I20-25, Z951, Z955, K40-46, K49, K50, and K75) from their linked electronic health records. The healthy controls were participants who reported to be non-smoking with a BMI < 30 and had no diagnoses of other cardiovascular diseases and known comorbidities, such as epilepsy and arrhythmias.

### Sample Quality Control (QC)

Sample QC measures that were provided by the UKB[24] (provided in square brackets are their FieldIDs from the UKB AMS) were also used to filter our cohort. Samples were retained that (i) had matching reported and genetic sex [22001], (ii) were not carriers of full or mosaic sex chromosome aneuploidies [22019], (iii) were not related to other UKB participants to the third degree [22021], (iv) were genetically identified as white British [22006]. Further GWAS QC thresholds (i.e., call rates > 98% and +/-3 standard deviations of calculated heterozygosity rate mean) were included to filter out low sample quality.

After sample QC, there were 24,954 CAD cases with 25,858 controls. While there were more healthy controls available (*n* = 182,772), the ratio of CAD cases and controls were kept relatively even to control for inflation of test statistics when using PLINK2[3] for single-SNV logistic regression (sLR) and to prevent convergence issues with REGENIE[17].

### Variant Quality Control (QC)

The following thresholds on GraphTyper metrics were used to ensure that variants called in the pVCFs were of high-quality: AAScore > 0.15, ABHet > 0.175, ABHom < 0.9, PASS_ratio > 0.05, QD > 6, and QUAL >= 10. Structural variants such as indels were also removed and multi-allelic variants were split for ease in downstream analysis.

Further GWAS QC thresholds were used to make certain only reliable variants were included in subsequent analysis. This included Hardy-Weinberg equilibrium *P* < 1 x 10^−6^, minor allele frequency (MAF) < 1 x 10^−5^, and a call rate > 98%.

After variant QC, there were 6,392,685 single nucleotide variants (SNVs) in chromosome 9 and 1,958,444 SNVs in chromosome 21. Due to the available sample size after QC, we added a further MAF 0.1% filter to remove ultra-rare SNVs before running sLR, REGENIE, fine mapping and VariantSpark. At this more stringent MAF threshold, there were 597,370 SNVs and 187,146 SNVs in chromosomes 9 and 21 respectively.

### Single-SNV Logistic Regression (sLR) with PLINK

A sLR with CAD status as response and the QC SNVs were modelled using PLINK2. The covariates included in the model were age, sex, BMI, and the first 10 principal components. The principal components were calculated using the UKB imputed genotypes to get genome-wide coverage. Genome-wide significance was the accepted *P* < 5 x 10^−8^ while marginal significance was *P* < 1 x 10^−5^.

### Whole-Genome Regression with REGENIE[17]

Whole genome regression, employing the REGENIE method, was used as an alternative approach for testing SNV associations with the same sets of covariate and phenotype data as above. The REGENIE run was divided into two steps. Initially, ridge regression predictors (*j*) are constructed for each block of the genome by partitioning it into consecutive blocks of *B* SNVs. These predictors are then combined through cross-validation to yield a single optimal predictor for estimating phenotype values. Subsequently, this predictor is divided into 23 chromosome-specific predictions for a leave-one-chromosome-out (LOCO) approach which is effective in preserving the power to detect associations by preventing proximal contamination[25,26]. The second step of REGENIE tests the association of each SNV by conditioning on the LOCO predictors, which are used as covariates to account for underlying background polygenic effects. The fast Firth logistic regression method provided by REGENIE was used for association testing, which improves the accuracy of logistic regression models by minimising bias in maximum-likelihood estimates through penalised likelihood. This approach ensures well-calibrated Type 1 errors and provides reliable estimates of SNV effect sizes and standard errors, especially when rare variants are being tested in the SNV set.

While both steps were available as applets in the RAP tool library, the initial step was conducted on our internal high-performance computing (HPC) environment using the previously downloaded imputed array genotype dataset to mitigate costs on the RAP. This approach was taken assuming that the predicted phenotype values would consistently capture the background genome-wide polygenic effect on the phenotype. Specifically, the genome-wide imputed array SNVs (with MAF > 0.01, missingness < 0.02, Het > 3, and GBR individuals) were employed to develop the regression predictors. The parameters included *B* = 500, *j* = 5 (default shrinkage parameter value = [0.01 0.25 0.5 0.75 0.99]), and 5-fold cross-validation to select the most effective predictors.

The LOCO predictions were then exported from the HPC to the RAP for the second step where the WGS QC’d SNVs on chromosomes 9 and 21 were tested using the fast Firth approximation implementation. Genome-wide significance was the accepted *P* < 5 x 10^−8^ while marginal significance was *P* < 1 x 10^−5^.

### Machine Learning Analysis with VariantSpark

VariantSpark is an Apache Spark-based application for building genome-wide random forest models and is not yet included in the RAP tool library. Following the guidelines available at https://documentation.dnanexus.com/developer/apps/developing-spark-apps, VariantSpark’s executable was created as a custom applet to run on RAP’s Spark cluster cloud architecture within the project space. To resolve conflicts between JAR file versions, VariantSpark needed to be compiled within the startup script rather than at applet build time, sacrificing some initialisation performance to ensure compatibility.

Smaller instance sizes are unable to handle the Spark application, which necessitates the use of more compute intensive and expensive instances for worker nodes. Because RAP does not currently differentiate between worker nodes and the primary node when specifying instance types, this necessarily increases the cost associated with the now underutilised driver node. As such, specifications allowing for the sizes of worker and driver nodes to be optimised separately by matching node capabilities more closely to their operational roles will result in better resource management and cost-efficiency.

The hyperparameters of the random forest models for both chromosomes 9 and 21 were *no_of_tree* (number of trees in forest) = 10K, *min_node_size* (minimum number of samples in node to be processed) = 10K, and *mtry_fraction* (fraction of variants evaluated at each node of a tree) = 0.1. The importance of a variable in the model was assessed using average Gini impurity measure.

The VariantSpark analysis of chromosome 9 was run on eight on-demand ‘mem2_ssd2_v2_x48’ instances which completed in 13 hours and 16 minutes, costing £179.31. In comparison, the VariantSpark analysis of chromosome 21 used six on-demand ‘mem2_ssd2_v2_x32’ instances which took 8 hours and 19 minutes to complete and cost £56.14.

### Annotating Association Results with Annovar[27]

Annovar was used to annotate association results from sLR, REGENIE, and VariantSpark. Namely, mapping SNVs to dbSNP rsIDs, genes and genetic function from UCSC’s RefSeq, and previous studies from the GWAS Catalog.

### Fine Mapping with PolyFun (SuSIE and FineMap)

A set of 5,179 SNVs at the associated *CDKN2B* locus, including a 500Kb flanking region, was subjected to functionally informed fine mapping using PolyFun[28] incorporating both the SuSie[10] and FINEMAP[9] methods. The approach adhered to the standard PolyFun workflow, integrating prior causal probabilities based on per-SNV heritability and summary LD information pre-calculated using individuals of European ancestry from the UK Biobank cohorts.

As the pre-calculated information was based on the hg19/GRCh37 reference, liftOver[29] was used to map the sLR summary statistics from hg38/GRCh38 to hg19/GRCh37. Following this, variants without pre-computed values were filtered out, leaving 4,843 remaining variants. For both methods, a maximum of 10 causal SNVs was specified for the locus. The locus spanned four overlapping LD regions, with variants assigned to region-specific causal sets.

Note that in the absence of a PolyFun applet on the RAP, the summary statistics file generated via sLR was downloaded from the RAP, and fine mapping was performed on an internal HPC environment.

## Acknowledgements

The analyses were conducted on the Research Analysis Platform (https://ukbiobank.dnanexus.com). We would like to thank and acknowledge DNAnexus and Amazon Web Services for their administrative support in enabling this study.

## Author Contributions

L.M.F.S, N.A.T., and D.C.B conceived the study. L.M.F.S, A.K., M.J.O., and B.H designed the study, analysed the data, and interpreted the results. L.M.F.S, A.K., and M.J.O. wrote the first draft. All authors reviewed, commented on, and approved the final version of the manuscript.

## Data Availability

Access to the UK Biobank dataset is upon application as described at http://www.ukbiobank.ac.uk/using-the-resource and with permission of UKB’s Research Ethics Committee. This study was conducted under the approved application 27483.

## Code Availability

Scripts to run RAPpoet, our scalable orchestration engine on DNAnexus CLI for RAP, is available at h<u>ttps://github.com/aehrc/RAPpoet</u>.

## Competing Interests Statement

All authors have no competing interests to declare.

## Figure Legends

**Figure 6.** A Driver-Worker Approach (RAPpoet) for Managing the Configuration of Requested Instances on UK Biobank RAP and for Parallelising the Processing of Files on Cloud Instances. The driver script, which remains on the user’s local machine, includes configurable environmental variables, such as the type of instance requested and number of batches to process in each instance. Meanwhile, the worker script, located in the RAP project environment, contains the actual commands to be executed on the instance that can be set by the user depending on their requirements.

**Figure 7.** Miami Plot of Chromosome 9 for single-SNV Logistic Regression (Top) and VariantSpark (Bottom). The locus 9p21.3 reached genome-wide significance in the logistic regression analysis and is the top association from VariantSpark. Note that the Gini score (bottom *y*-axis) is used to measure variable importance and while it is analogous to the traditional P-values of logistic regression to rank associations relative to other associations, it does not include multiple testing and does not have guiding significance thresholds.

**Figure 8.** Scatterplot of SNVs in the 9p21.3 locus with Posterior Inclusion Probability (PIP) (y-axis) against Chromosome 9 Position (x-axis) for FINEMAP (A) and SuSie (B). The first credible set of both approaches include the same 13 SNVs which are highlighted in dark purple and annotated with *rs*IDs.

**Figure 9.** Miami Plot of Chromosome 21 for Logistic Regression (Top) and REGENIE (Bottom). The locus 21q22.11, which has been previously associated with CAD, is only showing marginal association in the single-SNV logistic regression analysis. Conversely, a single SNV in the locus reached genome-wide significance in the REGENIE analysis.

**Figure 10.** Schematic of Pipeline for UK Biobank 500K Whole Genome Sequencing Dataset on RAP. RAPpoet was used to facilitate this pipeline on the UKB RAP.

## Notes

### Competing Interest Statement

The authors have declared no competing interest.

### Funding Statement

This study did not receive any funding

### Author Declarations

IRB of the Commonwealth Scientific and Industrial Research Organisation gave ethical approval for this work

### Summary of Updates

Table 1 added to main text. Results subsections shuffled to emphasis main focus of manuscript.

