## Supplementary Materials for "Optimizing UK Biobank Cloud Based Research Analysis Platform to Fine Map Coronary Artery Disease Loci in Whole Genome Sequencing Data"

**S1 Detailed Costings and Runtime of Pipeline**

Initially, all jobs were run as low-priority tasks to minimise costs, as low-priority tasks are restarted on 'spot' instances. However, at the time, low-priority tasks were simply terminated. Thus, jobs were set to normal-priority instead which were frequently restarted on 'on-demand' instances inflating costs unnecessarily. This issue has since been resolved by DNAnexus.

<u>Chromosome 21</u>

As outlined in Methods, the first step of our pipeline included sample filtering for the CAD cohort, QC filtering to ensure high quality variant calls, and the splitting of multi-allelic variants. RAPpoet facilitated effective job monitoring and allowed us to stay within the RAP limit of 500 concurrent instances. Using dxFUSE to rapidly stream data onto cloud instances, the 1,867 pVCFs from chromosome 21 were split into batches for 140 (i.e., 14 jobs), running 70 pVCFs concurrently. With this setting, step one of the pipeline had an average runtime of 1.75 minutes and an average cost of £0.052 per pVCF (Supplementary Table S10).

The next step of the pipeline included further single nucleotide variant (SNV) QC measures such as minor allele frequency (MAF) and call rate filters. Although the resulting BCFs from step one were 90% smaller in size, we opted to conduct further filtering before merging into a single BCF, as this would still result in a very large file (~20TB). However, due to the pVCFs being curated by region, rather than size or variant number, some BCFs had no variants remaining after the second QC step, causing PLINK2 to error out and crash the instance. As a workaround, the BCFs resulting from step one of the pipeline were partially concatenated before running QC step 2. The settings for RAPpoet were 160 files in each instance, concatenating 80 BCFs concurrently. This spun up 12 instances and took an average of 2.04 minutes and an average cost of £0.003 (Supplementary Table S12).

This completed all QC steps and files were merged into a single PLINK2 format file before running sLR. Using a 'mem2_ss1_v2_x48' spot instance, PLINK2 took 1 minute to merge all QC'd files and took 3 minutes to complete sLR. Including loading the Swiss Army Knife (SAK) app and writing out files, the whole job took 6 minutes costing £0.0332.

For REGENIE, a 'mem2_ssd1_v2_x32' spot instance was used to index and convert the QC'd merged PLINK2 files to BGEN. This step took 6 minutes costing £0.0178. A 'mem2_ssd1_v2_x64' on-demand instance completed step two of REGENIE in 31 minutes costing £0.2342.

For VariantSpark, a 'mem2_ssd2_v2_x32' high priority instance was used, which ran for 499 minutes, costing £56.14.

<u>Chromosome 9</u>

Prompted by the much larger number of pVCFs in chromosome 9 compared to chromosome 21, we attempted to optimise compute resources in the first step of the pipeline. The larger 'mem2_ssd1_v2_x96' instance type was used to accommodate bigger batches of 384 files with 192 pVCFs running concurrently. However, the dxFUSE filesystem failed as too many API calls were made which throttled the system and killed the instances. Therefore, batch sizing and the number of pVCFs to concurrently process were experimented with to ensure compute was still optimised without overloading dxFUSE (Supplementary Table S14).

The workaround was the same parameters as chromosome 21 (i.e., batch size = 140, concurrent pVCFs = 70) but repeated four times to process all 6,106 pVCFs in chromosome 9 on an instance size with less vCPUs. With this set up, step one of the pipeline had an average runtime of 1.80 minutes and an average cost of £0.029 per pVCF (Supplementary Table S11).

Similarly, for step two, we used the same parameters of batch sizing and concurrent pVCFs as chromosome 21 (i.e., batch size = 160, concurrent pVCFs = 80). For processing of 6,106 BCFs, this step was run using 10 instances and repeated 4 times sequentially (i.e., 10 jobs ran and completed before the next 10 started). This had an average runtime of 1.22 minutes and an average cost of £0.0055 per pVCF (Supplementary Table S13).

For the sLR, a single 'mem2_ssd1_v2_x48' spot instance was used to run PLINK2 to both merge all QC'd files and complete sLR modelling, which took 2 minutes and 10 minutes respectively. Including loading SAK and writing out files, the whole job took 15 minutes costing £0.0783.

To run REGENIE, the merged QC'd PLINK2 format files were converted to BGEN and indexed using a 'mem2_ssd1_v2_x32' spot instance. This took 6 minutes costing £0.0178. Step two of REGENIE was run on a 'mem2_ssd1_v2_x64' on-demand instance and completed in 91 minutes costing £2.7492.

For VariantSpark, a 'mem2_ssd2_v2_x48' high priority instance was used, which ran for 796 minutes, costing £179.31.

**Supplementary Table S14. Trials to Optimise Batch Sizing and Concurrent Files for Chr9 Step One.**
The two parameters of Batch and -P (i.e., concurrent files) were experimented with to determine the optimal values where compute was saturated without overloading dxFUSE.

| Test | Instance | Batch | -P | Failed | Error |
|---|---|---|---|---|---|
| **1** | mem2_ssd1_v2_x96 | 382 | 192 | All | The machine running the job became unresponsive |
| **2** | mem2_ssd1_v2_x48 | 382 | 100 | All | The machine running the job became unresponsive + Error while running the command (please refer to the job log for more information). Warning: Out of memory error occurred during this job. |
| **3** | mem2_ssd1_v2_x48 | 382 | 70 | Partial | The machine running the job became unresponsive + Error while running the command (please refer to the job log for more information). Warning: Out of memory error occurred during this job. |
| **4** | mem1_ssd1_v2_x72 | 140 | 70 | All | Error while running the command (please refer to the job log for more information). Warning: Out of memory error occurred during this job. |
| **5** | mem2_ssd1_v2_x48 | 140 | 70 | 0 | NA |
